# Bubble-PAPR: Phase I clinical evaluation of an ‘in-house’ developed prototype powered air-purifying respirator for use by healthcare workers

**DOI:** 10.1101/2022.07.14.22277643

**Authors:** Brendan A McGrath, Cliff Shelton, Angela Gardner, Ruth Coleman, James Lynch, Peter G Alexander, Glen Cooper

## Abstract

**Objectives:** We aimed to design and produce a low-cost, ergonomic, hood-integrated Powered Air-Purifying Respirator (Bubble-PAPR) for pandemic healthcare use, offering optimal and equitable protection to all staff. We hypothesised that participants would rate Bubble-PAPR more highly than current FFP3 face mask respiratory protective equipment (RPE).

**Design:** Rapid design and evaluation cycles occurred based on the identified user needs. We conducted diary card and focus group exercises to identify relevant tasks requiring RPE. Lab-based safety standards established against British Standard BS-EN-12941 and EU2016/425. Questionnaire-based usability data from participating frontline healthcare staff before (usual RPE) and after using Bubble-PAPR.

**Setting:** Overseen by a trial safety committee, evaluation progressed sequentially through laboratory, simulated, low-risk, then high-risk clinical environments of a single tertiary NHS hospital.

**Participants:** 15 staff completed diary cards and focus groups. 91 staff from a range of clinical and non-clinical roles completed the study, wearing Bubble-PAPR for a median of 45 minutes (IQR 30-80 [15-120]). Participants self-reported a range of heights (mean 1.7m [SD 0.1, range 1.5-2.0]), weights (72.4kg [16.0, 47-127]) and body mass indices (25.3 [4.7,16.7-42.9]).

**Outcome measures:** Primary: “*How comfortable do you feel in your PPE?”* (Likert scale bounded by 1 [very uncomfortable] to 7 [very comfortable]). Secondary outcomes: perceived safety, communication, anxiety, discomfort, and performance.

**Results:** Bubble-PAPR mean comfort score was 5.64(SD 1.55) versus usual FFP3 2.96(1.44) (mean difference 2.68 (95% CI 2.23-3.14, p<0.001). There was a significant difference in favour of Bubble-PAPR across all secondary outcomes.

**Conclusions:** Bubble-PAPR achieved its primary purpose of keeping staff safe from airborne particulate material whilst improving comfort and the user experience. The design and development of Bubble-PAPR were conducted using a careful evaluation strategy addressing key regulatory and safety steps, in contrast to many devices rapidly developed and deployed during the pandemic.

**Trial registration:** IRAS ID:288493, REC Ref:21/WA/0018. ClinicalTrials.gov (NCT04681365).

**Strengths and limitations of this study:**

- We employed user-centred design, engineering optimisation and staged feasibility testing to develop a novel Powered Air-Purifying Respirator (Bubble-PAPR) for use specifically in frontline healthcare settings.
- Diverse, frontline healthcare staff compared Bubble-PAPR with usual FFP3 face masks.
- The design and development of Bubble-PAPR were conducted using a careful strategy addressing key regulatory and safety steps, in contrast to many devices rapidly developed and deployed during the pandemic.
- Bubble-PAPR is an excellent example of developing a cosmopolitan network that could become a key feature of future system resilience.

## Introduction

The COVID-19 global pandemic created a worldwide shortage of personal protective equipment (PPE)^1^ and highlighted significant usability issues in current PPE products.^2^ In addition to direct contact, airborne diseases may be spread by aerosol or droplet transmission. Aerosol transmission may be mitigated by the appropriate use of respiratory protective equipment (RPE), a particular classification of personal protective equipment (PPE). However, respiratory protective equipment is used as part of a hierarchy of control measures and is usually considered a last resort. This is because RPE only protects individual workers, is prone to failure or misuse (wrong RPE for the wrong task/environment) and wearers may get a false sense of security, encouraging risk-taking behaviours.^3^ A range of inspiratory filtering devices exist: dust masks, half-face masks, full-face masks and powered (fan-assisted) respirators. Powered respirators include: half/full-face masks, helmets, hoods and visors. Though not used in healthcare, for completeness, breathing apparatuses are systems that supply an independent, positive pressure supply of breathing-quality air.

Face masks may be classified by considering the level of protection they offer the wearer to inhalation of environmental contaminants. Simple surgical face masks or ‘nuisance’ dust masks do not entirely filter droplets or aerosols. Filtering face piece (FFP) masks comprise layers of synthetic non-woven material with interleaved filtration layers and provide protection against small airborne particles (aerosols). Different types and constructions of FFP masks can be classified by their ability to filter small particles. Particulate filters can be classified as low (P1) to high (P3) efficiency, filtering between 80% of particles smaller than 2 micrometres to 99.95% of particles smaller than 0.5 micrometres, respectively (Table 1).^4^ Respiratory protection can therefore be considered in terms of a combination of the filtering ability of the device relative to the exposure environment and its fit on the wearer’s face. A device is considered adequate if it has the capacity to reduce the wearer’s exposure to a hazardous substance to acceptable levels (to comply with occupational exposure limit values). Devices can be reusable, but the majority are single-use. Masks are difficult to recycle due to their layered construction and the pandemic contributed to an unprecedented rise in RPE-related clinical waste.^5^

**Table 1.**
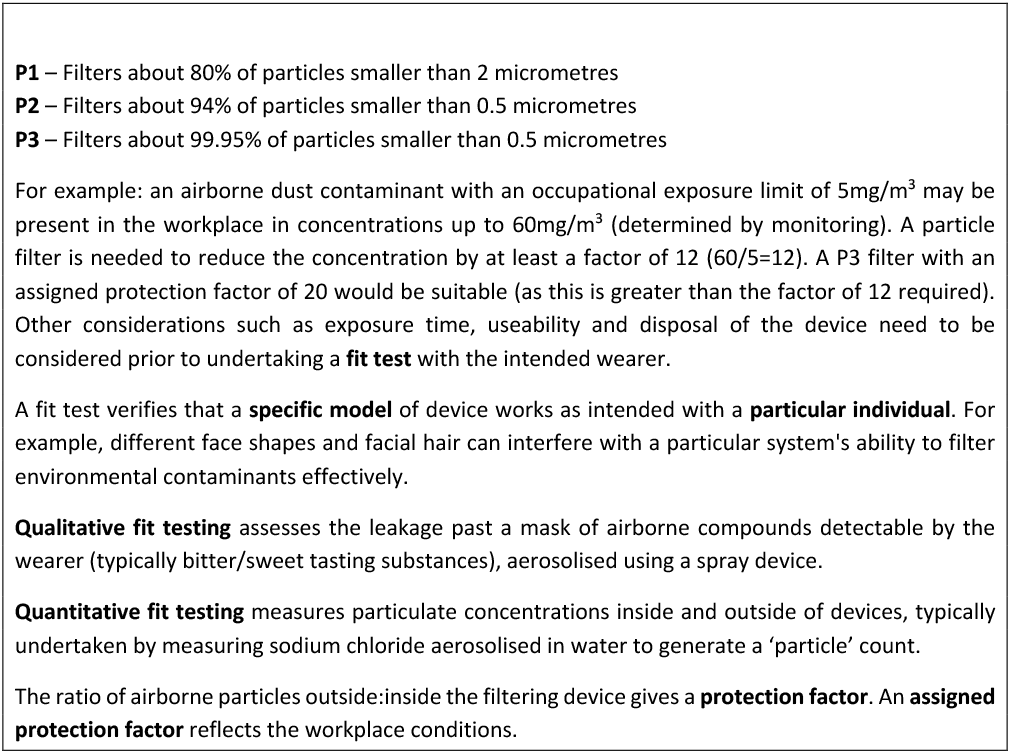
Classification of particulate filters, with a worked example and fit testing.

The majority of RPE used in healthcare settings are disposable face masks adopted from industry. Masks are not designed to be worn for long periods or repeated shifts, may restrict vision and communication, may cause facial damage due to their tight fit, and require multiple time-consuming ‘fit tests’ for each model of the device for each staff member. All these issues were highlighted in the context of the 2002-2004 SARS epidemic.^6^ More appropriate solutions for prolonged and repeated use include powered air-purifying respirators (PAPRs). But, again, these are not designed primarily for healthcare, are heavy, noisy, expensive, difficult to clean to clinical standards, and not suitable for the specific needs in frontline healthcare environments.

There have been several widely reported ‘homemade’ or ‘MacGyvered’ devices that well-intentioned groups or individuals developed to protect staff and patients during the pandemic.^7^ None of these devices sought or achieved independent certification or provided data to support safety.^8^ Turner and colleagues proposed a framework for the safer adoption of novel devices^7^ which: defines the problem and reviews existing solutions, benchmarks safety indices for the devices, and then evaluates it in a structured manner through simulated, low- and then high-risk clinical settings Table S1 (Supplemental). Broad stakeholder feedback is encouraged through iterative review cycles, re-design and improvements.

Considering the above, our project aimed to design and produce a low-cost, ergonomic, hood-integrated PAPR for use in frontline healthcare settings. Our objectives were to focus on user-centred design, engineering optimisation, staged feasibility testing, certification, intellectual property protection and then rapid manufacture and distribution. We also aimed to design the PAPR to be re-used, refurbished and recycled where possible, using readily available, simple and interchangeable key parts which proved difficult to source during the early stages of the pandemic. Finally, by designing an available, affordable PAPR system that could be cleaned appropriately and re-used between different staff, we aimed to provide equitable access to high-quality RPE that offered optimal protection to *all* staff, wherever they worked.^9^ We hypothesised that participants would rate Bubble-PAPR more highly than current FFP3 face mask RPE across the domains of comfort, perceived safety and communication.

## Methods

The design team brought together frontline clinical staff based in the Wythenshawe Hospital Acute Intensive Care Unit (ICU) of Manchester University NHS Foundation Trust (MFT), an experienced product design consultancy (Designing Science Limited, Middlesex, UK) and the technical expertise of the School of Engineering at the University of Manchester (UoM). Research Ethical and Health Research Authority approval (IRAS ID:288493, REC Ref:21/WA/0018) was granted. The study was sponsored by MFT, who acted as the manufacturer of this in-house prototype device, which became known as Bubble-PAPR. The study protocol, analysis plan and recruitment metrics were registered and reported at ClinicalTrials.gov (NCT04681365). User needs assessment was conducted through a series of workplace diary card exercises documenting typical activities undertaken by frontline healthcare staff, synthesised in focus groups. Staff were invited to participate (by email and posters in rest areas) from clinical locations where RPE was mandated within the hospital. The first two respondents from each area were recruited to the diary card and focus group activities. Rapid design and evaluation cycles occurred based on the identified user needs. In addition, evaluation of early prototypes occurred in simulated clinical environments, collecting usability data from participants.

### Patient and public involvement

Public and Patient involvement was undertaken through the Manchester Academic Critical Care research group’s patient forum. There were powerful accounts from patients who regularly described not being able to understand what hospital staff wearing PPE were saying and being troubled that they had no idea what their carers looked like. These reports led us to focus on prioritising the ease of communication with Bubble-PAPR. Staff participants who were invited to wear Bubble-PAPR were again recruited from clinical locations where RPE was mandated, by direct invitation from the research team. All relevant staff working in the area were approached until a maximum of six staff had been recruited per shift (the most that the research team could reasonably accommodate per shift), or the recruitment target had been met.

A Trial Safety Committee was established to oversee the results of laboratory and bench testing of the prototype, initial safety data, usability and adverse event data at each stage of the evaluation. The Committee met prior to commencing clinical evaluation. It was tasked with the decision to allow the evaluation to proceed between phases: simulated clinical environment, low-risk (non-infectious) clinical environment and high-risk clinical environment (COVID-19 wards and ICUs). A final iteration of Bubble-PAPR was further tested in high-risk environments. Prior to first use, several device safety checks were independently undertaken by the MFT Electrical and Biomedical Engineering Department and INSPEC International Ltd, Salford, UK). A short report based on the criteria detailed in Table S2 (Supplemental) was presented to the Trial Safety Committee. The first ten study participants to wear Bubble-PAPR underwent ‘fit testing’ with a particulometer (TSI Portacount Fit Tester 8040, TSI Instruments Ltd, Buckinghamshire, UK) following a standard protocol derived from the UK Government’s Health and Safety Executive.^12^ This INDG-479 protocol requires a ‘Fit Factor’ pass level of 100 for FFP3/N95 face masks and 500 for full face masks/hoods. European Conformity Standard EN12941 requires an applied fit factor of 40 for a ‘loose-fitting hood’ PAPR; the equivalent of a nominal protection factor of at least 500 (accepting an inward leakage of 0.2% with a P3 class filter. See Table 1). By comparison, the minimal fit factor for an FFP3 mask in a clinical environment is 100. Tests were conducted in an ICU side room with a particle generator to reach background counts between 70,000 to 100,000 particles/cm^3^.

The primary outcome was based on Davis’ technology acceptance model (perceived usefulness and perceived ease-of-use overcoming barriers to adoption)^10^. First, staff were asked to rate their experiences using current RPE (a variety of re-useable or disposable FFP3 masks) using a series of questions based on Likert-type scales (see Supplemental material). Next, safe use of the Bubble-PAPR was explained, and instructions for use were provided, supported by videos of donning, doffing, cleaning and storage. Bubble-PAPR was then worn during simulated/clinical use where the usual tasks (identified in the focus groups) were undertaken. Finally, after removal (doffing) of Bubble-PAPR, staff were immediately invited to complete a second questionnaire focused on the prototype. Free text comments were also invited.

The primary endpoint was staff rating of the comfort of Bubble-PAPR (versus current FFP3 face masks). Secondary endpoints focused on communication and perceived safety. Specifically, this was staff ratings of the prototype in terms of: how safe participants felt, ease of communication with colleagues, and ease of communication with patients (again, Bubble-PAPR versus current FFP3 face masks). In parallel, in-house device feasibility testing was conducted in the hospital environment to test ergonomics and air particle filtration. We tested against existing conformity standards for PAPRs relevant at the time of development (British Standard BS EN 12941 [Respiratory Protective Devices: Powered filtering devices incorporating a helmet or hood] and the European Union Personal Protective Equipment Directive EU2016/425). ^4 11^

A pilot evaluation was conducted in August 2020 to test the questionnaires and to assess the likely population means for the test scores (Table S3 Supplemental). We calculated a sample size of 20 participants would be required for each phase of the evaluation to detect a significant difference between usual PPE and Bubble-PAPR, based on a mean difference of 2.5 (SD 0.9) points on the 7-point Likert scale identified during the pilot evaluation (alpha = 0.05, 90% power). In addition, we allowed for a 5% dropout and missing data rate, concluding 22 participants per phase. All variables were explored via appropriate graphical and descriptive statistics for completeness and form. Analyses were conducted in RStudio 2020 (Boston, MA, www.rstudio.com). Analyses were performed separately for each phase for presentation to the Trial Safety Committee, with a pooled analysis conducted at the study conclusion. Comparisons between groups (current RPE vs Bubble-PAPR) were made using a paired t-test or Wilcoxon signed-rank test as appropriate.

## Results

The final design of Bubble-PAPR is shown schematically in Figure 1 (www.bubble-papr.com). The device safety checks and fit testing results are presented in Tables S2, S3 and S4 (Supplemental), respectively, demonstrating a mean fit factor of 16,961. Additional particulometer tests were undertaken with deliberate tears up to 20 cm in the hood using a dummy head. The lowest fit factor recorded with the damaged hood was 1,123. Therefore, the Trial Safety Committee concluded that the Bubble-PAPR performed its primary purpose of adequately protecting staff from airborne environmental contaminants.

**Figure 1.**
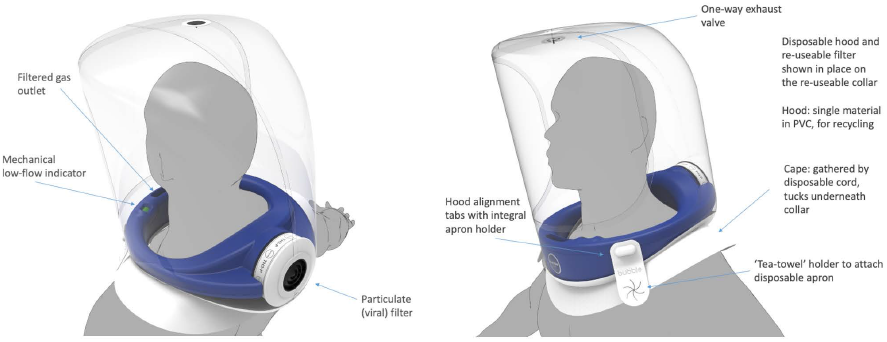
Bubble-PAPR comprises a medical-grade foam neck collar and a separate PVC hood. The universal fit collar draws air in through a filter via an impellor powered by an external battery. The collar has a mechanical low flow indicator and can be cleaned and reused by different users. The semi-rigid hood is pulled over the collar before donning and is secured by integrated straps.

Fifteen staff contributed to the diary and focus group exercises, generating a list of tasks to be undertaken. One staff member from the 16 invited could not attend the focus group meeting. Ninety-one staff wore Bubble-PAPR for a median of 45 (IQR 30-90, range 10-150) minutes between 3^rd^ March and 21^st^ December 2021. No staff who were approached during their clinical shifts were unwilling or unable to trial Bubble-PAPR. There were no Bubble-PAPR-related safety incidents reported during the study. Staff undertook all clinical duties identified by the focus groups and diary card exercise, either in the simulation suite (n=22) or clinical settings (n=22 low-risk, n=25 high-risk, n=22 high-risk with final iteration). Participants predominantly declared as female (69%), and were from a range of clinical and non-clinical roles, Figure S1 (Supplemental). Staff self-reported a range of heights (mean 1.7m [SD 0.1, range 1.5-2.0]), weights (72.4kg [16.0, 47-127]) and body mass indices (25.3 [4.7,16.7-42.9]), Figure S2 (Supplemental). Fifty-two percent of participants reported that they normally wore glasses, with 31% wearing glasses during the evaluation. All participants described at least 6 month’s experience with FFP3 face masks on a regular basis (“most shifts”), with a combination of re-useable (typically 3M™ 6000 Series Respirators) and single use (typically 3M™ Aura™ 9330 or equivalent) face masks. No participants described using PAPRs in the six months prior to recruitment. All participants completed all mandatory questionnaire sections.

With pooled data for the primary outcome, “*How comfortable do you feel in your PPE?”* (Likert scale bounded by 1 [very uncomfortable] to 7 [very comfortable]), Bubble-PAPR mean score was 5.64 (SD 1.55) versus usual FFP3 face mask 2.96 (1.44), Figure 2. There was a mean difference of 2.68 (95% CI 2.23-3.14, p<0.001). Secondary outcomes focused on communication and perceived safety. For the question, “*How safe do you feel in your PPE?*”, Bubble-PAPR mean score was 6.15 (0.94) vs usual FFP3 face mask 5.43 (0.98); mean difference 0.73 (95% CI 0.45-1.00, p<0.001), Figure 2. Figure 3 demonstrates communication outcomes for all 91 comparisons of Bubble-PAPR versus usual FFP3 face masks. All adjusted comparisons were significant (p<0.001) in favour of Bubble-PAPR for communicating with both colleagues and patients (Table 2).

**Table 2.** Rating scales, summary results and comparisons across the questionnaire domains.

|  | PPE | Q1 Confidence in donning | Q2 Confidence in donning without dislodging other PPE | Q3 Wear with glasses/goggles | Q4 Protect yourself from respiratory infection | Q5 Protect patient from infection from you | Q6 Safely care for your patient | Q7 Safely roll patient | Q8 Speak to staff | Q9 Be heard by staff | Q10 Speak to patient | Q11 Be heard by patient | Q12 Doff safely | Q13 Doff without dislodging glasses |
| --- | --- | --- | --- | --- | --- | --- | --- | --- | --- | --- | --- | --- | --- | --- |
| Rating scale | From: | 0 - no confidence | 0 - no confidence | 0 - no confidence | 0 - no confidence | 0 - no confidence | 0 - no confidence | 0 - no confidence | 0 - no confidence | 0 - no confidence | 0 - no confidence | 0 - no confidence | 0 - no confidence | 0 - no confidence |
|  | To: | 10 - fully confident | 10 - fully confident | 10 - fully confident | 10 - fully confident | 10 - fully confident | 10 - fully confident | 10 - fully confident | 10 - fully confident | 10 - fully confident | 10 - fully confident | 10 - fully confident | 10 - fully confident | 10 - fully confident |
| RPE type | FFP3 | 8.9 (1.4) [3 - 10] | 8.3 (2) [2 - 10] | 6.9 (2.6) [2 - 10] | 8.2 (1.6) [4 - 10] | 8.2 (1.7) [2 - 10] | 8.4 (1.4) [5 - 10] | 8.2 (1.8) [2 - 10] | 5.1 (2.4) [1 - 10] | 4.9 (2.3) [1 - 10] | 4.8 (2.4) [1 - 10] | 4.7 (2.5) [1 - 10] | 8.1 (1.9) [2 - 10] | 6.2 (2.5) [1 - 10] |
|  | Bubble | 7.4 (1.8) [3 - 10] | 7.7 (1.8) [2 - 10] | 7.6 (1.9) [3 - 10] | 8.6 (1.6) [3 - 10] | 8.5 (1.8) [2 - 10] | 8.0 (2) [2 - 10] | 7.8 (2.2) [2 - 10] | 7.5 (2.4) [1 - 10] | 7.1 (2.3) [1 - 10] | 7.8 (2.1) [2 - 10] | 7.4 (2.4) [1 - 10] | 8.0 (1.8) [2 - 10] | 7.8 (1.9) [2 - 10] |
| Comparison | Mean difference | -1.48 | -0.55 | 0.7 | 0.43 | 0.3 | -0.42 | -0.42 | 2.38 | 2.16 | 2.99 | 2.7 | -0.1 | 1.66 |
|  | 95% CI | -1.9 to -0.99 | -1.12 to 0.02 | -0.00 to 1.40 | -0.04 to 0.89 | -0.18 to 0.78 | -0.91 to 0.07 | -0.98 to 0.15 | 1.66 to 3.11 | 1.45 to 2.88 | 2.36 to 3.62 | 1.97 to 3.433 | -0.63 to 0.43 | 0.98 to 2.34 |
|  |  | Favours FFP3 | No difference | Favours Bubble | No difference | No difference | No difference | No difference | Favours Bubble | Favours Bubble | Favours Bubble | Favours Bubble | No difference | Favours Bubble |
|  | Adjusted p | <0.001 | 0.058 | 0.049 | 0.070 | 0.217 | 0.092 | 0.144 | <0.001 | <0.001 | <0.001 | <0.001 | 0.711 | <0.001 |

|  | PPE | Q14 How safe does it feel | Q15 Worried about own health | Q16 Worried others health | Q17 Comfortable | Q18 Don ease | Q19 Doff ease | Q20a Restricted communication | Q20b Vision distorted with Bubble | Q20c Read monitors, computers, and notes with Bubble | Q21 Pressure marks on head/face | Q22 Pain on head/face | Q23 Leave clinical area early due to RPE |
| --- | --- | --- | --- | --- | --- | --- | --- | --- | --- | --- | --- | --- | --- |
| Rating scale | From: | 1 - very unsafe | 1 - not worried at all | 1 - not worried at all | 1 - very uncomfortable | 1 - not at all easy | 1 - not at all easy | 1 - not at all restricted | 1 - not at all affected | 1 - clear at all times | 1 - never | 1 - never | 1 - never |
|  | To: | 7 very safe | 7 - very worried | 7 - very worried | 7 - very comfortable | 7 - very easy | 7 - very easy | 7 - very restricted | 7 - very affected | 7 - not clear at all | 7 - always | 7 - always | 7 - always |
| RPE type | FFP3 | 5.4 (1) [3 - 7] | 3.2 (1.5) [1 - 7] | 3.2 (1.5) [1 - 7] | 3 (1.4) [1 - 6] | 4.9 (1.3) [2 - 7] | 5.1 (1.3) [2 - 7] | 5.4 (1.4) [1 - 7] | - | - | 5.8 (1.4) [1 - 7] | 5.3 (1.4) [1 - 7] | 3.6 (1.6) [1 - 7] |
|  | Bubble | 6.2 (0.9) [3 - 7] | 2.3 (1.6) [1 - 7] | 2.3 (1.7) [1 - 7] | 5.6 (1.6) [1 - 7] | 5.5 (1.4) [2 - 7] | 5.7 (1.2) [2 - 7] | 3.9 (1.7) [1 - 7] | 3.2 (1.9) [1 - 7] | 5.6 (1.5) [2 - 7] | 1.3 (0.8) [1 - 6] | 1.4 (0.9) [1 - 6] | 1.5 (1) [1 - 6] |
| Comparison | Mean difference | 0.73 | -0.92 | -0.93 | 2.68 | 0.62 | 0.63 | -1.49 | - | - | -4.54 | -3.99 | -2.13 |
|  | 95% CI | 0.45 to 0.99 | -1.36 to -0.49 | -1.36 to -0.48 | 2.23 to 3.14 | 0.21 to 1.02 | 0.26 to 0.99 | -1.95 to -1.04 | - | - | -4.90 to -4.17 | -4.35 to -3.63 | -2.51 to -1.75 |
|  |  | Favours Bubble | Favours Bubble | Favours Bubble | Favours Bubble | Favours Bubble | Favours Bubble | Favours Bubble | No comparator | No comparator | Favours Bubble | Favours Bubble | Favours Bubble |
|  | Adjusted p | <0.001 | <0.001 | <0.001 | <0.001 | 0.003 | 0.002 | <0.001 | - | - | <0.001 | <0.001 | <0.001 |

**Figure 2.**
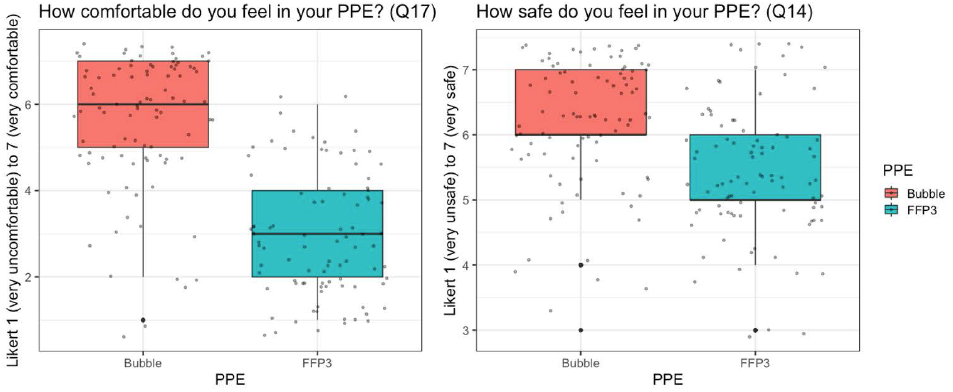
Reported comfort (primary) and safety (secondary) outcomes for Bubble-PAPR vs usual FFP3 face masks.

**Figure 3.**
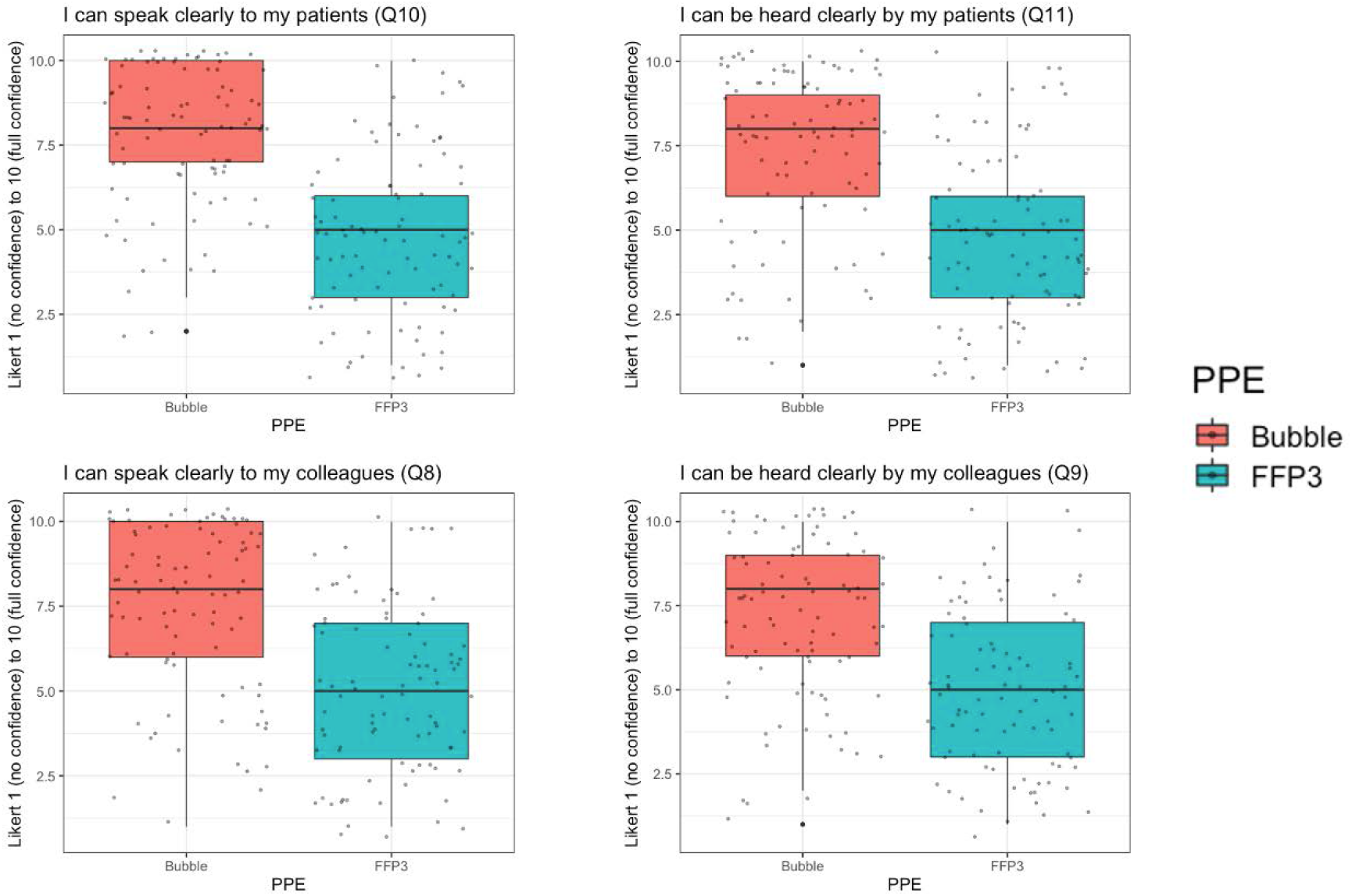
Secondary communication outcomes where a higher Likert scale response was considered better.

Secondary outcomes where a lower Likert response was considered better are presented in Figure S3 (Supplemental). These focussed on whether staff were worried about themselves or others whilst wearing RPE, whether the devices caused pressure or pain or if communication was impaired. Finally, staff were asked if they had to cut short a clinical (or simulated) encounter due to discomfort with their RPE. Again, there was a significant difference in favour of Bubble-PAPR for all metrics (all p<0.001, Table 2).

During the initial phases, there was no significant difference between staff reporting ease of donning and doffing of Bubble-PAPR and usual PPE (which staff had used for many months at the time of the evaluation). However, pooled results saw staff becoming more familiar with the Bubble, and Bubble-PAPR was rated easier to don and doff when compared with usual FFP3 face masks (adjusted p=0.003 and 0.002 respectively), Table 2 and Figure S4 (Supplemental). Free text comments were reviewed and categorised into positive, negative and neutral comments (Figures S5-7 Supplemental). Most comments focused on the noise of the device, which improved throughout the project as the impellor and motor were made quieter in later design iterations.

## Discussion

Our project developed an innovative prototype PAPR explicitly designed for prolonged healthcare use in high-risk clinical environments. Bubble-PAPR achieved its primary purpose of protecting staff from airborne potentially infectious material whilst also being rated significantly higher for comfort (the primary outcome), perceived safety, and communication with colleagues and patients (secondary outcomes) than usual RPE. Bubble-PAPR was used in all relevant simulated and clinical scenarios identified by detailed staff diary cards, making the results of this study extremely relevant to hospital-based healthcare workers.

Bubble-PAPR was rapidly developed based on the lived experiences of frontline staff during the early stages of the coronavirus pandemic, addressing the unmet needs of reliable, high-quality, universal and available RPE with improved comfort and communication when compared to usual FFP3 face masks. Staff overwhelmingly recognised the importance of facial visualisation when communicating with colleagues and patients. When combined with the improved comfort of wearing a PAPR over usual RPE, participants rated Bubble-PAPR consistently highly across all comparator domains.

This relatively simple evaluation study was preceded by a rapid design and prototyping phase, producing a working prototype within a few weeks. Despite the speed and agility demonstrated by the design team, we adhered to relevant standards, following a tiered evaluation within the governance structure of an approved and regulated research project. Bubble-PAPR was only introduced into higher-risk environments following review by the Trial Safety Committee. This approach contrasted with many rapidly developed or adopted RPE systems that became prevalent during the pandemic, often disseminated via social media and almost always without any meaningful safety or useability data.^7, 13, 14^ Whilst the PPE shortages experienced during the pandemic drove many of these innovations and adaptations, we recognised the importance of a methodical approach to design, development and testing of our prototype, both in the laboratory and clinical settings. We recommend others to follow the framework proposed by Duggan et al. for the development of novel medical devices, with regular reviews of safety and useability data within the framework of a robust and transparent clinical trial.^7^

Our study has some limitations. The design of Bubble-PAPR addressed many of the issues identified by the same staff who subsequently evaluated the prototype. Whilst our study protocol allowed evaluation only within our Trust owing to the ‘in-house’ manufacturing exemption for testing, it is not unreasonable to expect similar results if our prototype were evaluated elsewhere. Although this may be considered a weakness of the study, many of the shortcomings of the PPE provided to frontline health workers around the world are well described and are essentially the same as those identified in our project.^15, 16^ Furthermore, we evaluated Bubble-PAPR against single-use and reusable FFP3 face masks, which could be construed as comparing two different classes of RPE. However, Bubble-PAPR was designed and developed to provide a viable alternative to FFP3 class face masks, in contrast to the more usual healthcare use of PAPRs – selectively available on a limited basis to specific users or groups. Our detailed analysis of work diary cards from various clinical staff ensured that Bubble-PAPR was used for all relevant procedures undertaken by medical, nursing, healthcare assistant, allied healthcare professional (speech and language therapy, physiotherapy, pharmacy), administrative anddomestic staff in the clinical area. This perspective is unique within respiratory protective equipment product evaluation studies.^17, 18^

Our study did not directly evaluate the patient experience with staff wearing different RPE. However, the patient experience was reflected in the user specifications identified around communication, and anecdotal feedback was positive from patients, especially around facial visibility and verbal and non-verbal communication. In addition, when contrasted with FFP3 face masks, speech and language therapists reported that demonstrating speech and swallow exercises was suddenly possible with Bubble-PAPR and that the transparent nature of the hood overcame the communication barriers that can be so devastating for those with hearing impairments.^19^ Although designed to be potentially recyclable, future work should address the environmental impact of PVC hoods with reusable collars compared to single-use or reusable FFP3 face masks.

## Conclusions

Our study has demonstrated that Bubble-PAPR achieved its primary purpose of keeping staff safe from airborne particulate material whilst improving comfort, communication and the user experience. It is likely that the patient experience was also enhanced. Bubble-PAPR has been patented (PCT/GB2021/052147) and subsequently licenced to a UK Healthcare company for large-scale manufacture and distribution to frontline NHS and other workers. The pandemic drove unprecedented collaboration between clinicians, academics and industry. Bubble-PAPR is an excellent example of developing a cosmopolitan network that could become a key feature of future system resilience. The design and development of Bubble-PAPR were conducted using a careful strategy addressing key regulatory and safety steps, in contrast to many devices rapidly developed and deployed during the pandemic.

## Data Availability

The full dataset is potentially commercially sensitive. However, all data produced in the present study are available upon reasonable request to the authors

## Acknowledgements

We are grateful to our funders (detailed below) for supporting this project and the staff who participated in the evaluation. In addition, we are indebted to the designers, engineers and staff who gave their time freely during the early stages of the coronavirus pandemic to work tirelessly on designing, building, testing and refining Bubble-PAPR. Specifically:

Patrick Hall, Designing Science Ltd (www.designingscience.co.uk)

Andrew Spragg, Industrial design consultant

Andrew Forbes, XK Design Ltd

James Corden, Manchester University Hospital NHS Foundation Trust Innovation Team

Nick Duggan, Innovation Consultant, Zuas (www.zuas.io)

GAMA Healthcare Ltd, Hemel Hempstead, UK (www.gamahealthcare.com)

## Data availability statement

Due to the commercial sensitivity of the intellectual property licensed at the conclusion of this project, the full dataset is not publicly available. However, the corresponding author will consider requests to disclose the dataset on an individual basis if necessary.

## Trial registration

Research Ethical and Health Research Authority approval (IRAS ID:288493, REC Ref:21/WA/0018). Manchester University NHS Foundation Trust sponsored the study, acting as the manufacturer of this in-house prototype device. The study protocol, analysis plan and recruitment metrics were registered and reported at ClinicalTrials.gov (NCT04681365).

## Author contributions

All authors critically revised the manuscript for important intellectual content and approved the final manuscript. BAM attests that all listed authors meet authorship criteria and that no others meeting the criteria have been omitted.

**BAM:** conception and design, collection, analysis and interpretation of data, drafting and revision of the manuscript, and final approval of the version to be published. Participant recruitment.

**CS:** qualitative work package conception and design, analysis and interpretation of data. Participant recruitment. Drafting and revision of the final manuscript, and final approval of the version to be published.

**AG:** qualitative work package design, analysis and interpretation of data. Drafting and revision of the final manuscript, and final approval of the version to be published.

**RC:** design, collection, and interpretation of data, drafting and revision of the manuscript, and final approval of the version to be published.

**JL:** design, collection, and interpretation of data, drafting and revision of the manuscript, and final approval of the version to be published.

**PGA:** design, collection, and interpretation of data, drafting and revision of the manuscript, and final approval of the version to be published.

**GC:** design, collection, and interpretation of data, drafting and revision of the manuscript, and final approval of the version to be published. Manufacturing and engineering lead.

## Provenance and peer review

Not commissioned; externally peer reviewed.

## Ethics approval

Research Ethical and Health Research Authority approval (IRAS ID:288493, REC Ref:21/WA/0018) was granted from Wales REC5 on 27^th^ January 2021. The study was sponsored by Manchester University NHS Foundation Trust, who acted as the manufacturer of this in-house prototype device. The study protocol, analysis plan and recruitment metrics were registered and reported at ClinicalTrials.gov (NCT04681365).

## Table and Figure legends

**Table S1. (Supplemental)** Proposed framework for the safer adoption of a MacGyvered device. Adapted from Turner and colleagues.^7^

1. Define the problem and rule out the suitability of existing solutions
2. List benchmark safety indices for the device
3. Seek broader feedback from all stakeholders on the design’s utility and potential pitfalls.
4. Perform laboratory-based and in situ simulations.
5. Introduce into low-risk clinical settings after local due process and patient consent.
6. Introduce into higher-risk clinical settings with a discrete group of trained ‘super-users’.
7. Encourage an iterative cycle of feedback, review, re-design and improvement.
8. **Do not:** adopt, publish, endorse or disseminate via social media a MacGyvered device without data to support safety.

**Table S2.**
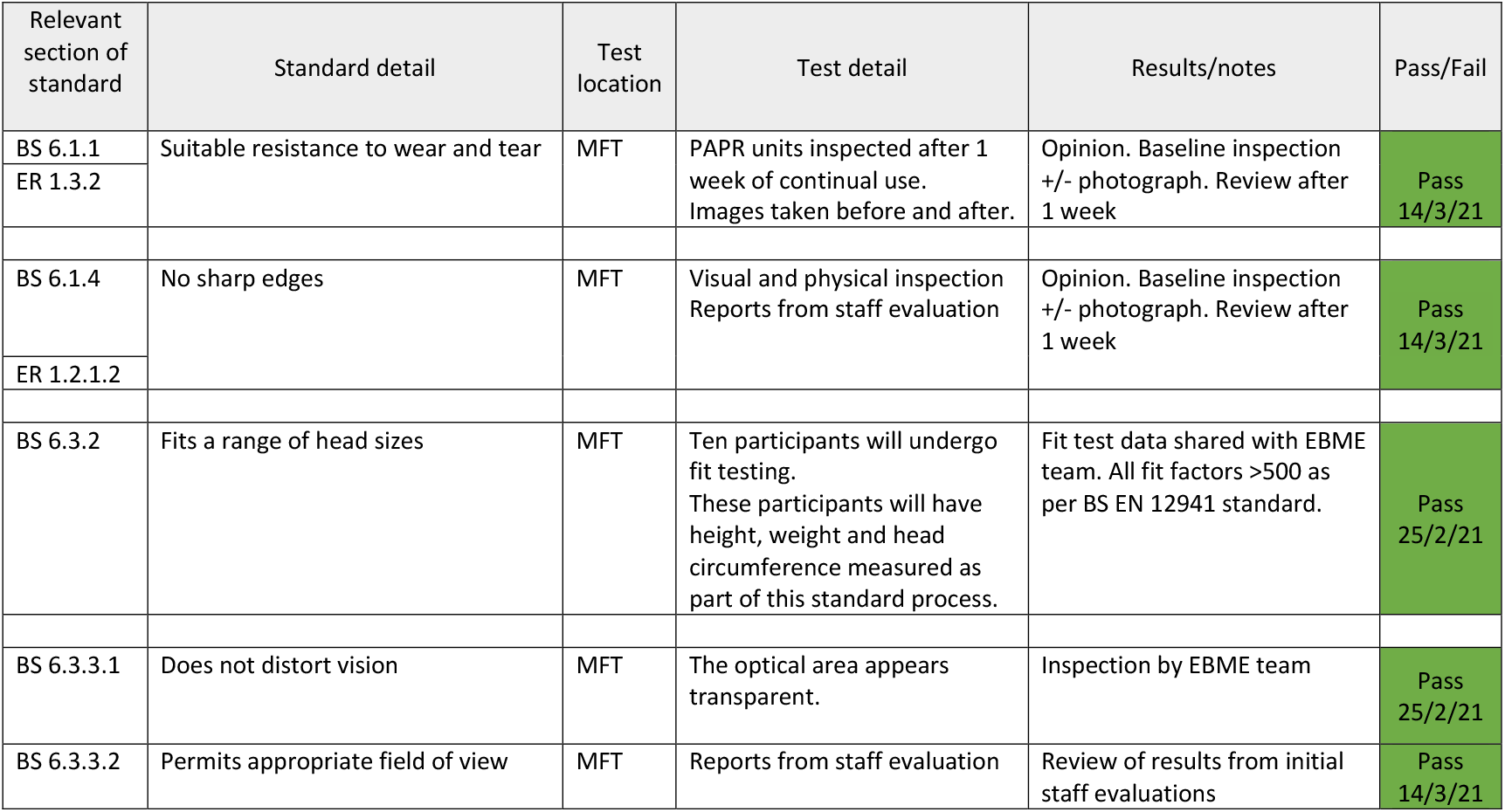
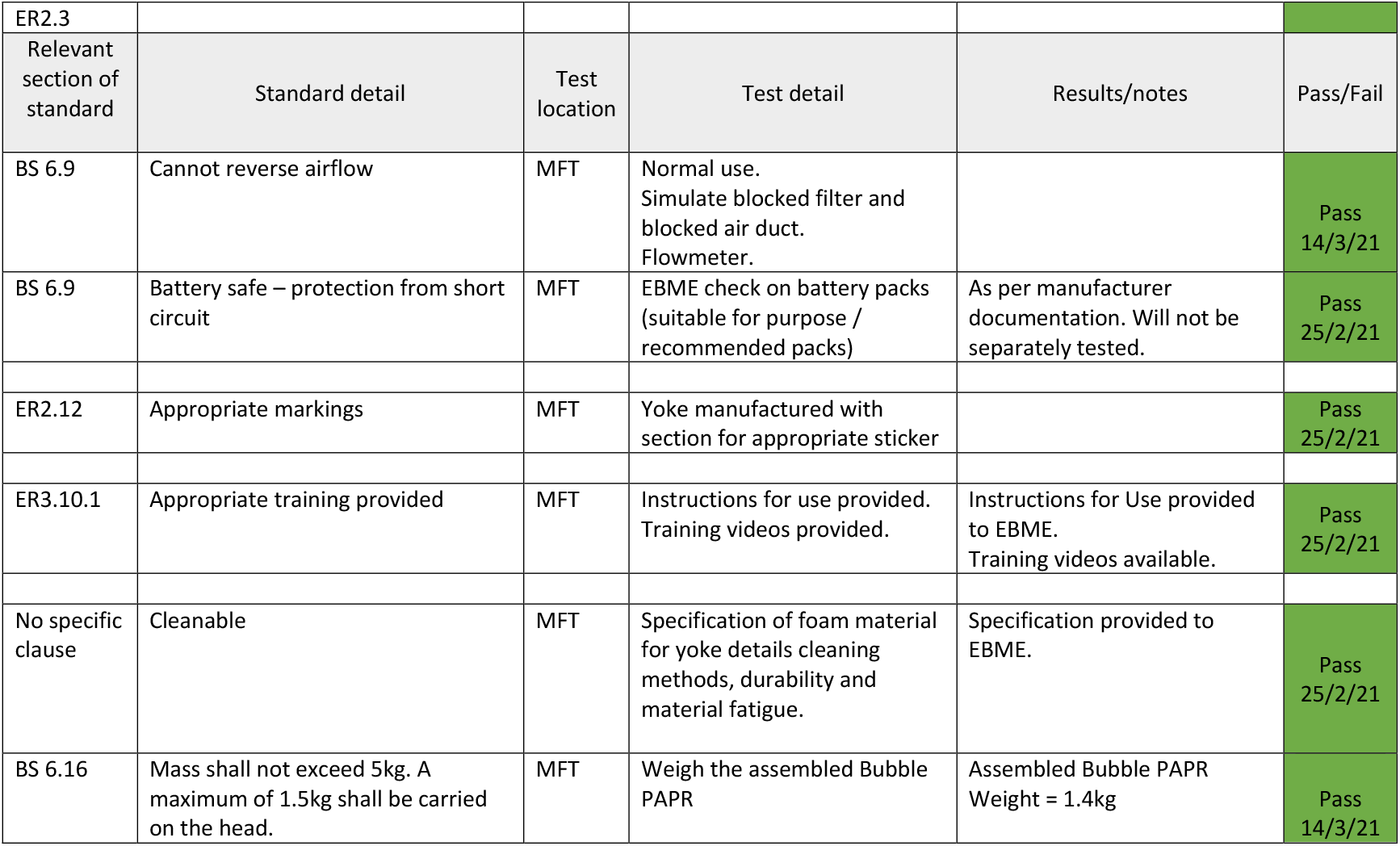
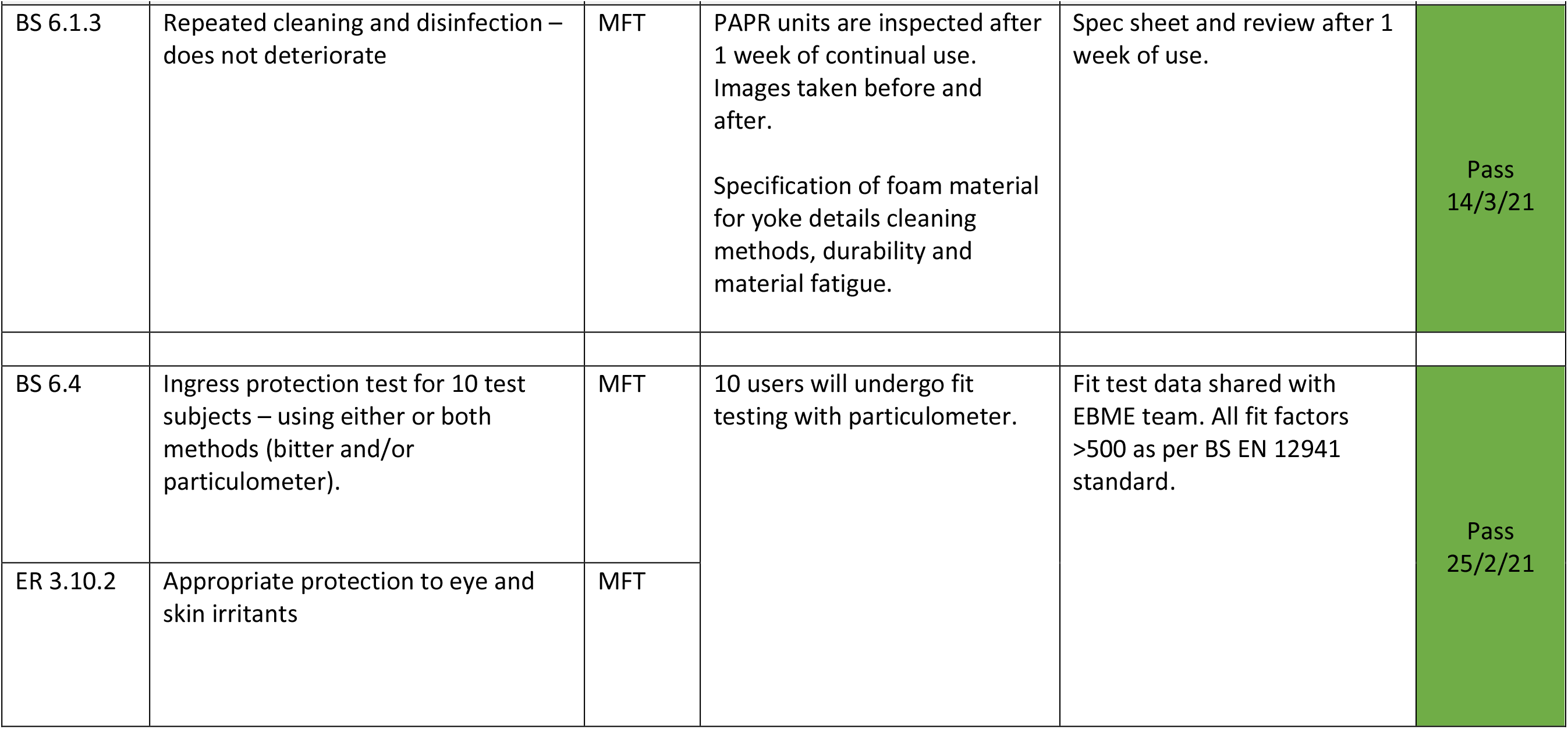
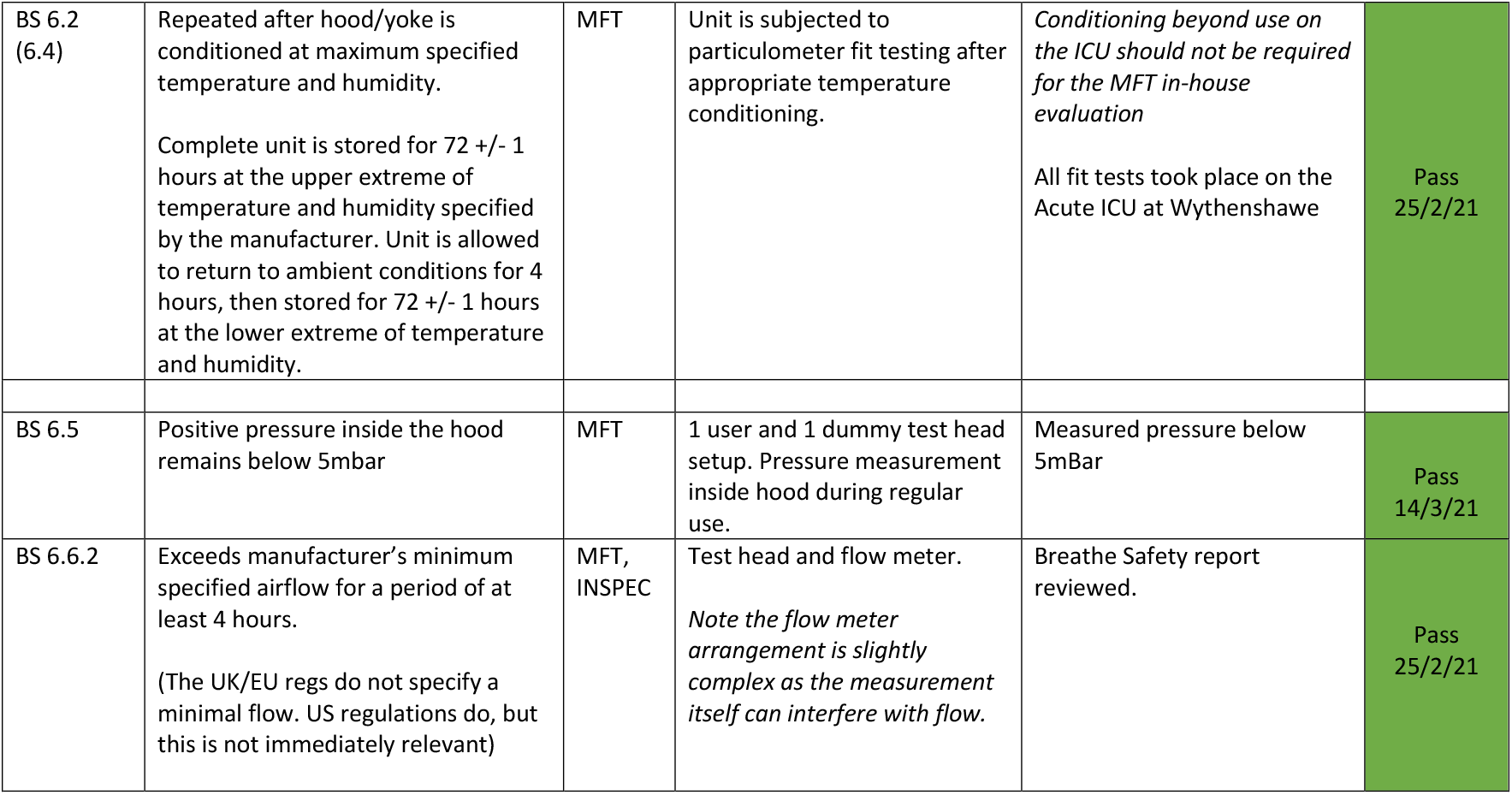
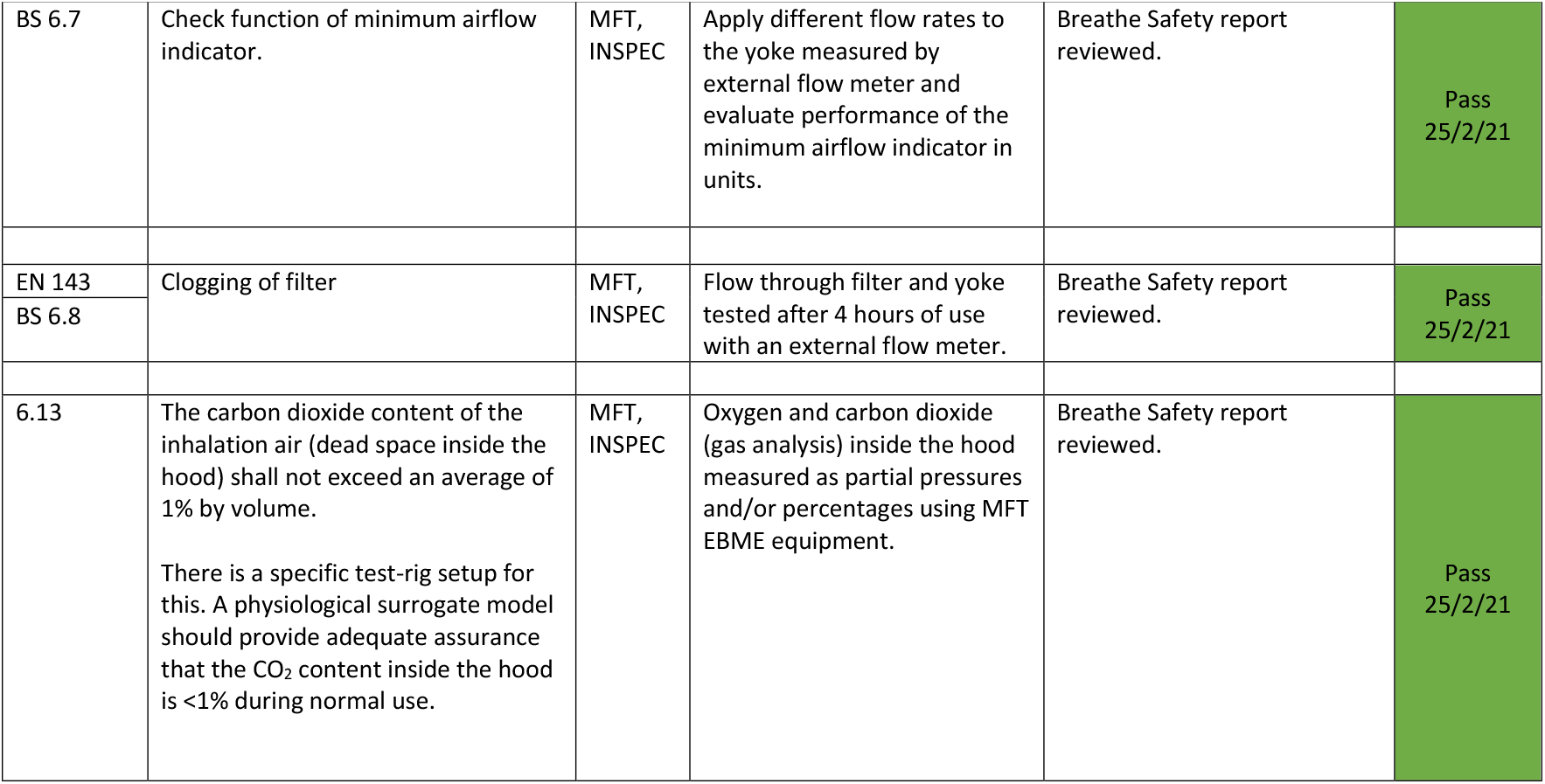
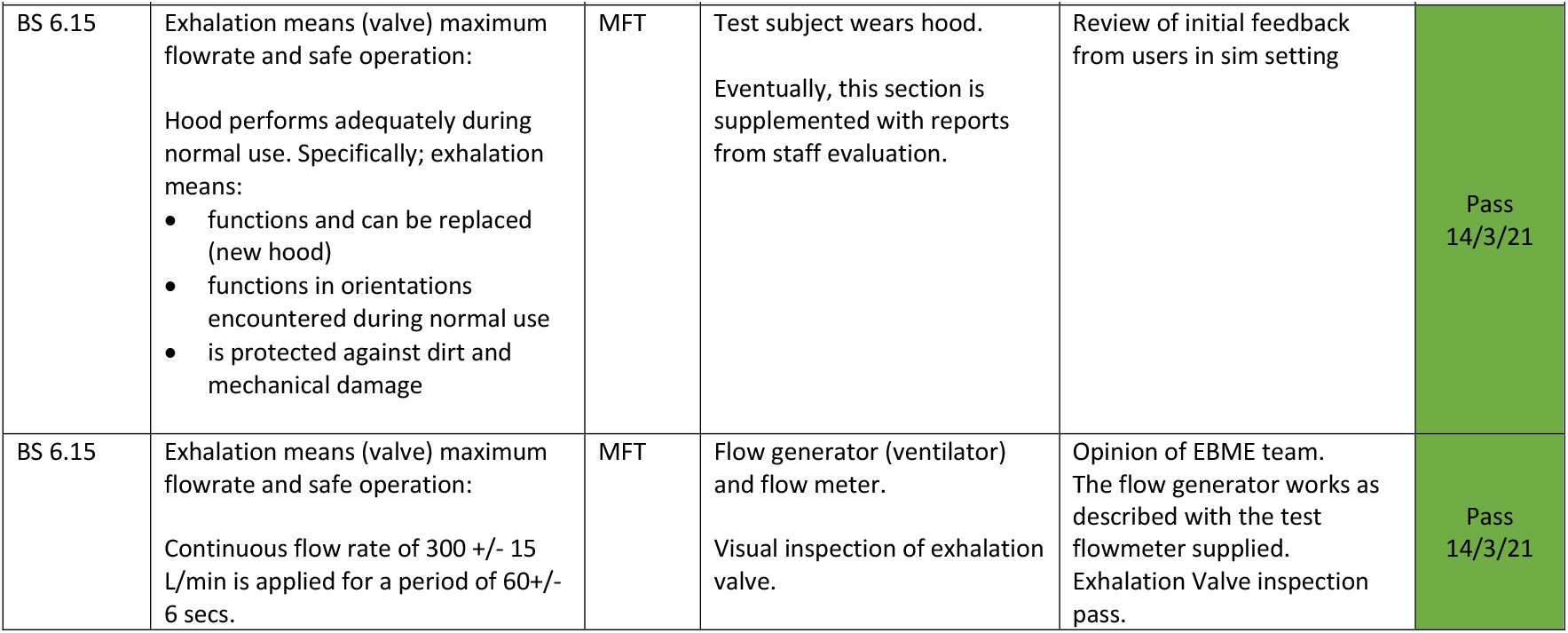
(Supplemental). Lab-based testing of the Bubble PAPR prior to clinical evaluation. All of the bench tests detailed below were carried out by the Electrical and Biomedical Engineering (EMBE) team based at Wythenshawe Hospital (MFT), the University of Manchester Mechanical Aerospace and Civil Engineering team (UoM) or by INSPEC International, Salford, UK. A PAPR unit was supplied, and the Instructions for Use were followed by the independent tester, with a judgement made if they fulfilled particular requirements. Some requirements are supplemented by the qualitative or quantitative data collected in the questionnaires. Standards used were British Standard EN12941 (BS, 2008) and the European Regulations for Respiratory Protective Equipment EU2016/425 (ER, 2016).

| Relevant section of standard | Standard detail | Test location | Test detail | Results/notes | Pass/Fail |
| --- | --- | --- | --- | --- | --- |
| BS 6.1.1 | Suitable resistance to wear and tear | MFT | PAPR units inspected after 1 week of continual use. Images taken before and after. | Opinion. Baseline inspection +/- photograph. Review after 1 week | Pass<br>14/3/21 |
| ER 1.3.2 |  |  |  |  |  |
| BS 6.1.4 | No sharp edges | MFT | Visual and physical inspection<br>Reports from staff evaluation | Opinion. Baseline inspection +/- photograph. Review after 1 week | Pass<br>14/3/21 |
| ER 1.2.1.2 |  |  |  |  |  |
| BS 6.3.2 | Fits a range of head sizes | MFT | Ten participants will undergo fit testing.<br>These participants will have height, weight and head circumference measured as part of this standard process. | Fit test data shared with EBME team. All fit factors >500 as per BS EN 12941 standard. | Pass<br>25/2/21 |
| BS 6.3.3.1 | Does not distort vision | MFT | The optical area appears transparent. | Inspection by EBME team | Pass<br>25/2/21 |
| BS 6.3.3.2 | Permits appropriate field of view | MFT | Reports from staff evaluation | Review of results from initial staff evaluations | Pass<br>14/3/21 |

| ER2.3 |  |  |  |  |  |
| --- | --- | --- | --- | --- | --- |
| Relevant section of standard | Standard detail | Test location | Test detail | Results/notes | Pass/Fail |
| BS 6.9 | Cannot reverse airflow | MFT | Normal use.<br>Simulate blocked filter and blocked air duct.<br>Flowmeter. |  | Pass<br>14/3/21 |
| BS 6.9 | Battery safe – protection from short circuit | MFT | EBME check on battery packs (suitable for purpose / recommended packs) | As per manufacturer documentation. Will not be separately tested. | Pass<br>25/2/21 |
| ER2.12 | Appropriate markings | MFT | Yoke manufactured with section for appropriate sticker |  | Pass<br>25/2/21 |
| ER3.10.1 | Appropriate training provided | MFT | Instructions for use provided.<br>Training videos provided. | Instructions for Use provided to EBME.<br>Training videos available. | Pass<br>25/2/21 |
| No specific clause | Cleanable | MFT | Specification of foam material for yoke details cleaning methods, durability and material fatigue. | Specification provided to EBME. | Pass<br>25/2/21 |
| BS 6.16 | Mass shall not exceed 5kg. A maximum of 1.5kg shall be carried on the head. | MFT | Weigh the assembled Bubble PAPR | Assembled Bubble PAPR<br>Weight = 1.4kg | Pass<br>14/3/21 |
| BS 6.1.3 | Repeated cleaning and disinfection – does not deteriorate | MFT | PAPR units are inspected after 1 week of continual use. Images taken before and after.<br>Specification of foam material for yoke details cleaning methods, durability and material fatigue. | Spec sheet and review after 1 week of use. | Pass<br>14/3/21 |
| BS 6.4 | Ingress protection test for 10 test subjects – using either or both methods (bitter and/or particulometer). | MFT | 10 users will undergo fit testing with particulometer. | Fit test data shared with EBME team. All fit factors >500 as per BS EN 12941 standard. | Pass<br>25/2/21 |
| ER 3.10.2 | Appropriate protection to eye and skin irritants | MFT |  |  |  |
| BS 6.2 (6.4) | Repeated after hood/yoke is conditioned at maximum specified temperature and humidity.<br>Complete unit is stored for 72 +/- 1 hours at the upper extreme of temperature and humidity specified by the manufacturer. Unit is allowed to return to ambient conditions for 4 hours, then stored for 72 +/- 1 hours at the lower extreme of temperature and humidity. | MFT | Unit is subjected to particulometer fit testing after appropriate temperature conditioning. | *Conditioning beyond use on the ICU should not be required for the MFT in-house evaluation*<br>All fit tests took place on the Acute ICU at Wythenshawe | Pass<br>25/2/21 |
| BS 6.5 | Positive pressure inside the hood remains below 5mbar | MFT | 1 user and 1 dummy test head setup. Pressure measurement inside hood during regular use. | Measured pressure below 5mBar | Pass<br>14/3/21 |
| BS 6.6.2 | Exceeds manufacturer's minimum specified airflow for a period of at least 4 hours.<br>(The UK/EU regs do not specify a minimal flow. US regulations do, but this is not immediately relevant) | MFT, INSPEC | Test head and flow meter.<br>*Note the flow meter arrangement is slightly complex as the measurement itself can interfere with flow.* | Breathe Safety report reviewed. | Pass<br>25/2/21 |
| BS 6.7 | Check function of minimum airflow indicator. | MFT, INSPEC | Apply different flow rates to the yoke measured by external flow meter and evaluate performance of the minimum airflow indicator in units. | Breathe Safety report reviewed. | Pass<br>25/2/21 |
| EN 143 | Clogging of filter | MFT, INSPEC | Flow through filter and yoke tested after 4 hours of use with an external flow meter. | Breathe Safety report reviewed. | Pass<br>25/2/21 |
| BS 6.8 |  |  |  |  |  |
| 6.13 | The carbon dioxide content of the inhalation air (dead space inside the hood) shall not exceed an average of 1% by volume.<br>There is a specific test-rig setup for this. A physiological surrogate model should provide adequate assurance that the CO<sub>2</sub> content inside the hood is <1% during normal use. | MFT, INSPEC | Oxygen and carbon dioxide (gas analysis) inside the hood measured as partial pressures and/or percentages using MFT EBME equipment. | Breathe Safety report reviewed. | Pass<br>25/2/21 |
| BS 6.15 | Exhalation means (valve) maximum flowrate and safe operation:<br>Hood performs adequately during normal use. Specifically; exhalation means:<br>• functions and can be replaced (new hood)<br>• functions in orientations encountered during normal use<br>• is protected against dirt and mechanical damage | MFT | Test subject wears hood.<br>Eventually, this section is supplemented with reports from staff evaluation. | Review of initial feedback from users in sim setting | Pass<br>14/3/21 |
| BS 6.15 | Exhalation means (valve) maximum flowrate and safe operation:<br>Continuous flow rate of 300 +/- 15 L/min is applied for a period of 60+/- 6 secs. | MFT | Flow generator (ventilator) and flow meter.<br>Visual inspection of exhalation valve. | Opinion of EBME team. The flow generator works as described with the test flowmeter supplied. Exhalation Valve inspection pass. | Pass<br>14/3/21 |

**Table S3.**
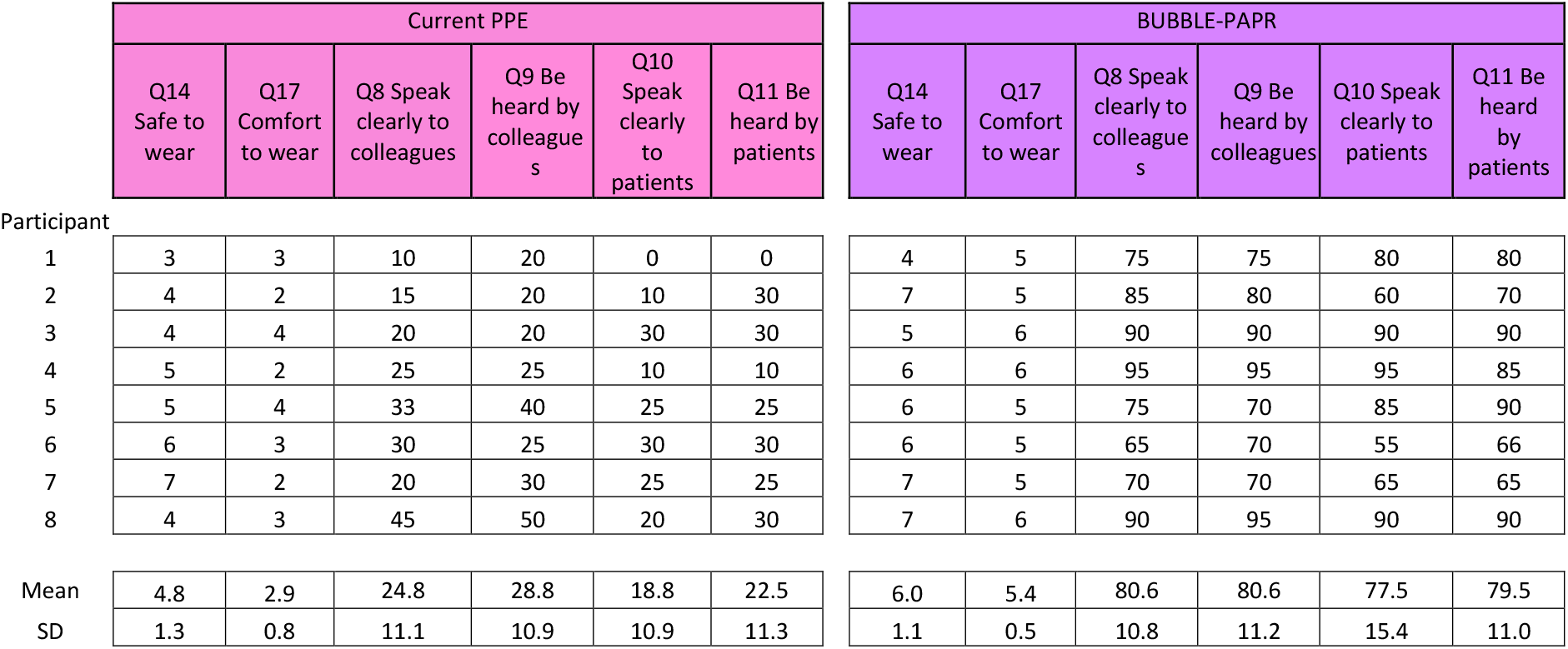
(Supplemental). Pilot data. Q14 & Q17 are Likert Scale items (rated 1-7) and Q8-11 are Visual Analogue Scale items (rated 0-100).

**Table S4.** (Supplemental) Fit testing data from the first 10 participants. Test protocol HSE INDG 479. Pass level set at a fit factor of 500.

| Subject | Self-reported height | Self-reported weight | BMI | Normal Breathing 1 | Deep Breathing | Head Side to Side | Head Up and Down | Talking | Bending at the waist | Normal Breathing 2 | Overall Fit Factor |
| --- | --- | --- | --- | --- | --- | --- | --- | --- | --- | --- | --- |
| **1** | 1.86 | 74 | 21.4 | 79705 | 43647 | 125478 | 11125 | 107899 | 1339 | 76152 | **7757** |
| **2** | 1.95 | 75 | 19.7 | 53792 | 52343 | 59440 | 51673 | 52733 | 50433 | 45961 | **52075** |
| **3** | NR | NR | NR | 38867 | 36699 | 37097 | 41474 | 39500 | 36884 | 37465 | **38217** |
| **4** | 1.82 | 65 | 19.6 | 17745 | 6622 | 3149 | 5028 | 31996 | 30520 | 31326 | **8539** |
| **5** | 1.65 | 55 | 20.2 | 24945 | 25215 | 3885 | 8097 | 28877 | 29107 | 24393 | **12268** |
| **6** | 1.67 | 58 | 20.8 | 24617 | 25608 | 25581 | 25225 | 20107 | 1924 | 23517 | **9088** |
| **7** | 1.52 | 47 | 20.3 | 28747 | 30700 | 33203 | 15275 | 31671 | 8327 | 26041 | **19829** |
| **8** | 1.65 | 65 | 23.9 | 27282 | 31318 | 34900 | 9697 | 29093 | 3514 | 25770 | **12544** |
| **9** | 1.83 | 115 | 34.3 | 11182 | 1123 | 11028 | 24524 | 24692 | 2537 | 23704 | **4408** |
| **10** | 1.59 | 75 | 29.7 | 25760 | 3419 | 16125 | 2433 | 25523 | 1552 | 26154 | **4588** |
| **Mean values** |  |  |  | **33264** | **25669** | **34989** | **19455** | **39209** | **16614** | **34048** | **16931** |

**Figure S1.**
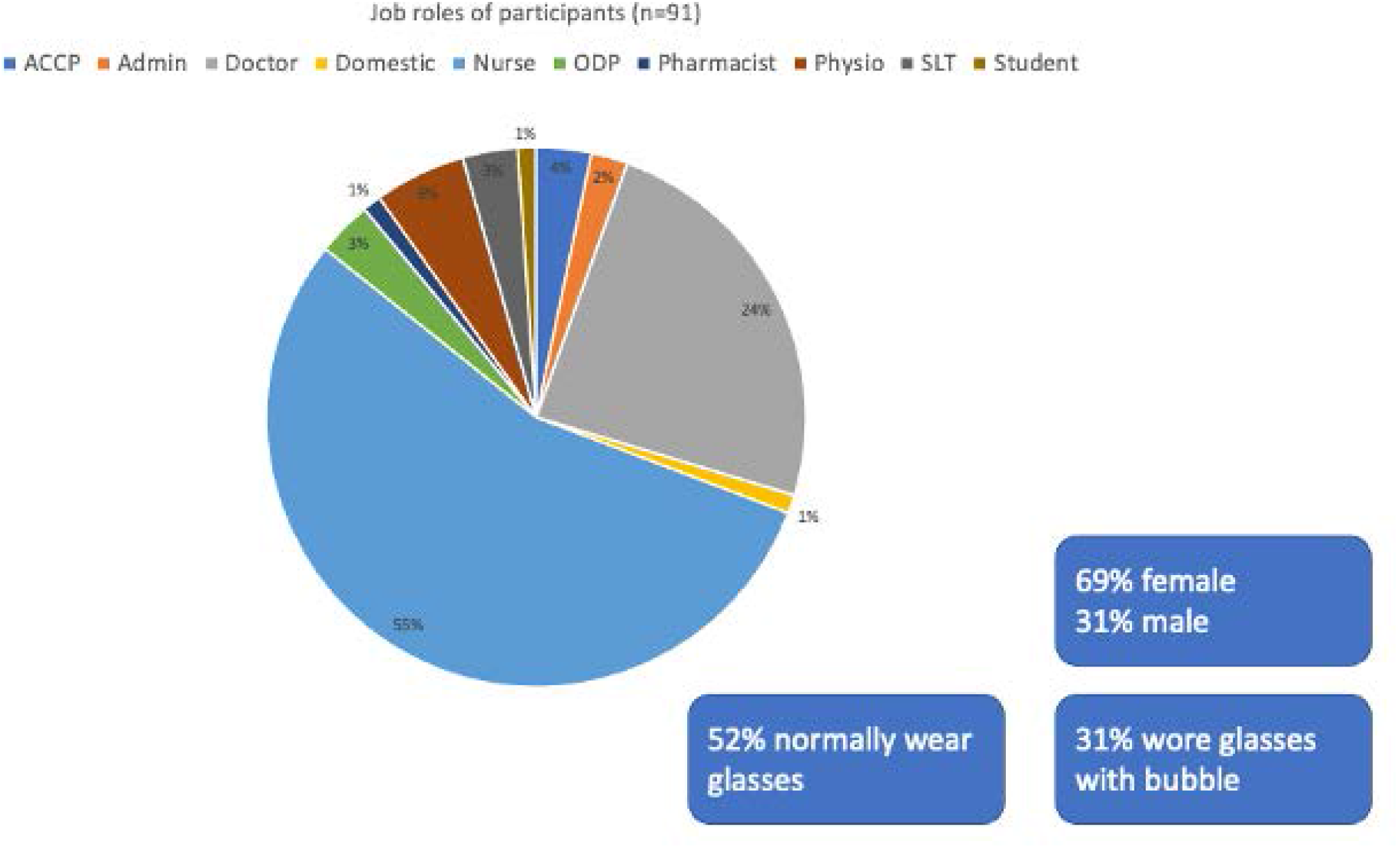
**(Supplemental)** Participant job roles.

**Figure S2.**
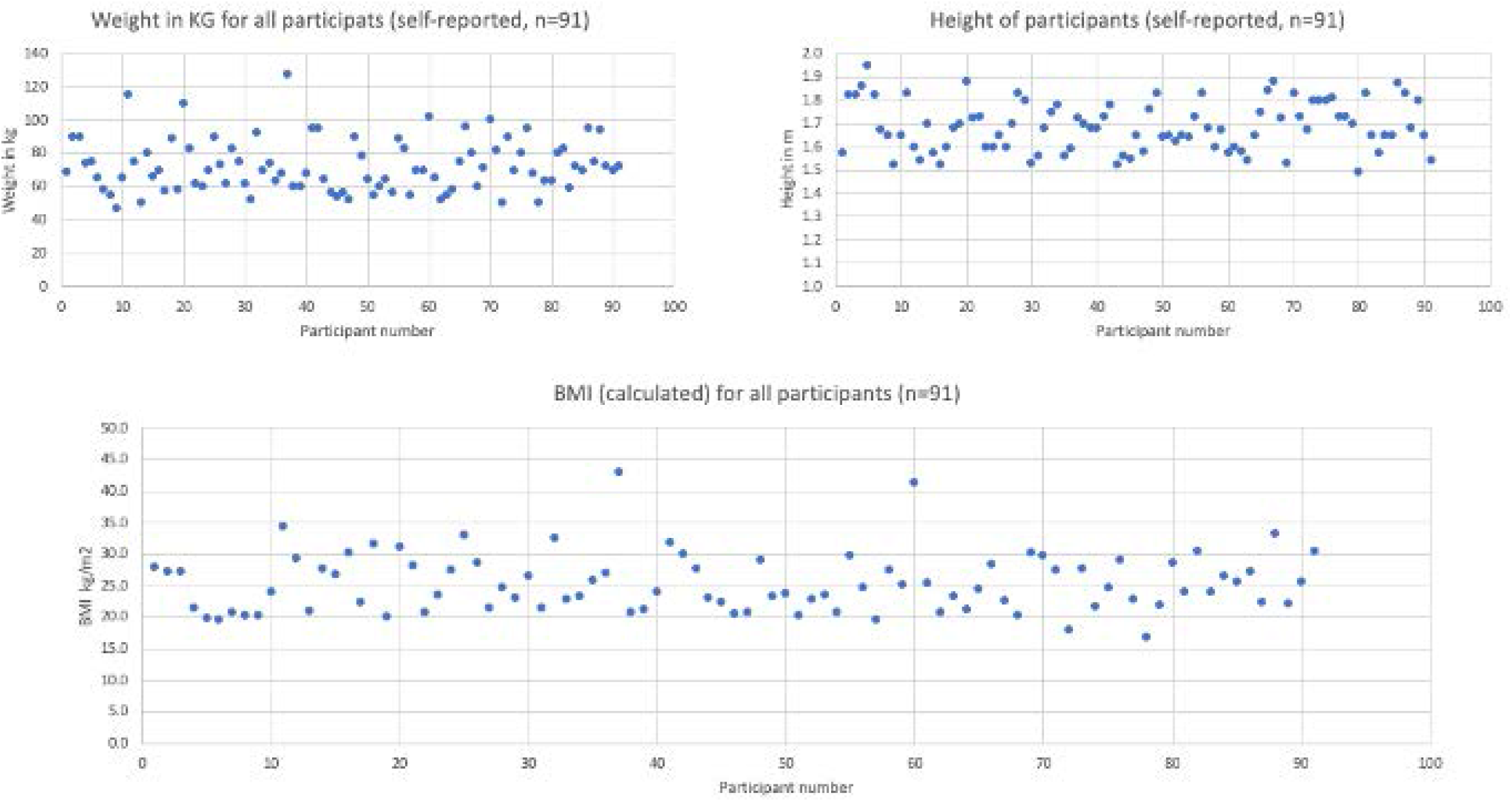
**(Supplemental)** Self-reported weight, height and BMI of staff participants.

**Figure S3.**
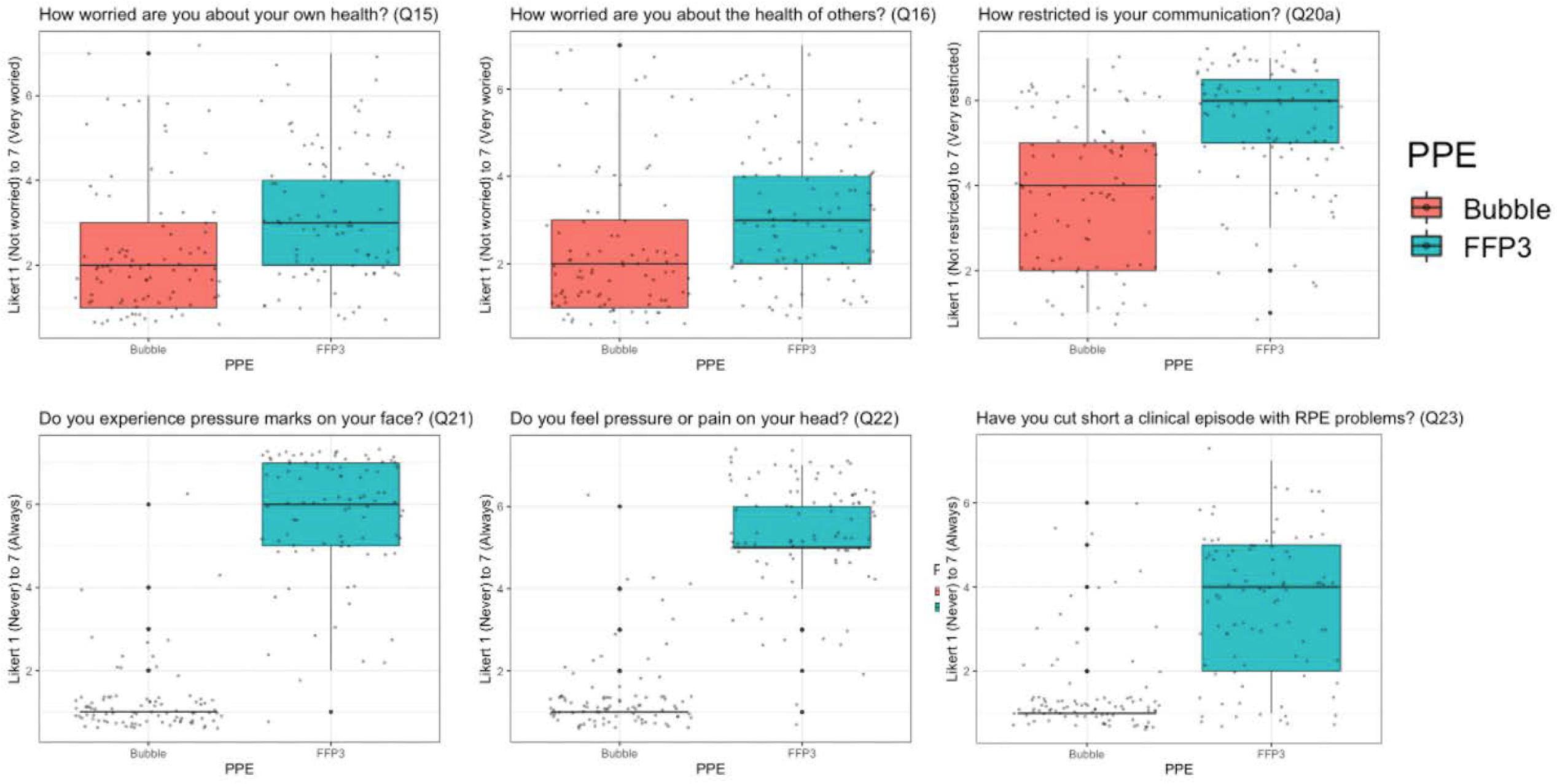
**(Supplemental)** Secondary outcomes where a lower Likert scale response was considered better.

**Figure S4.**
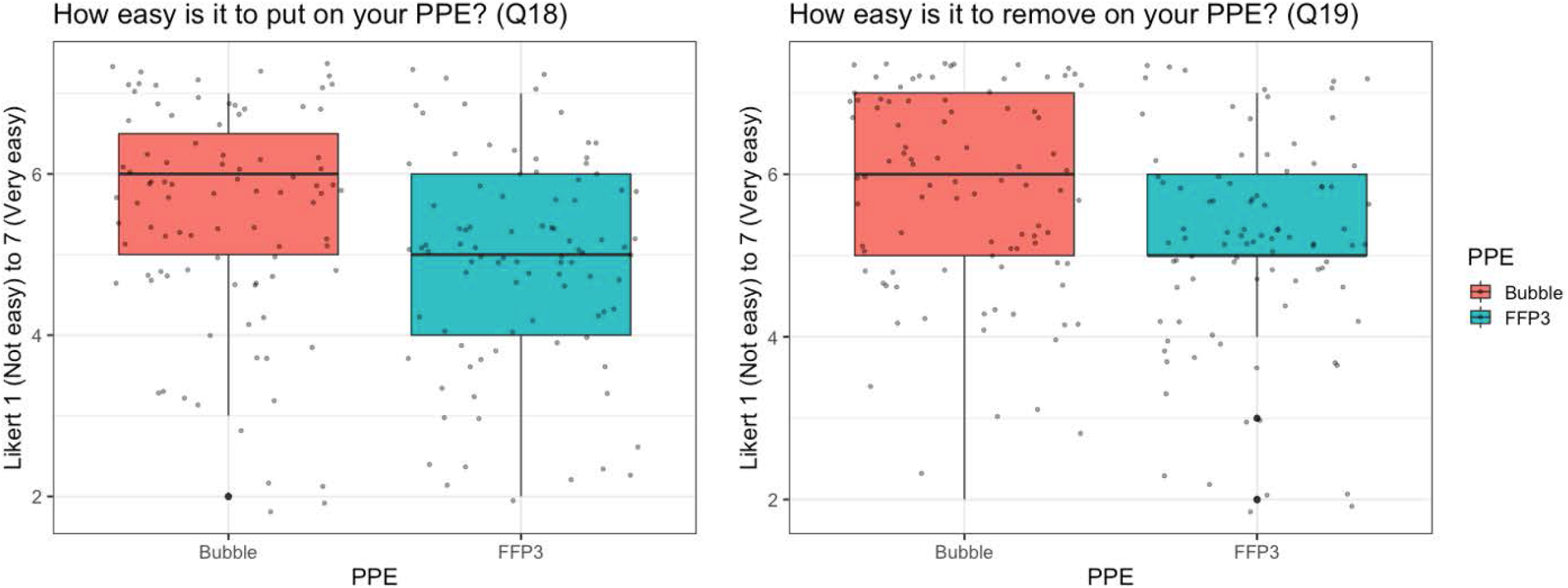
**(Supplemental)** Ease of donning and doffing of Respiratory Protective Equipment.

**Figure S5.**
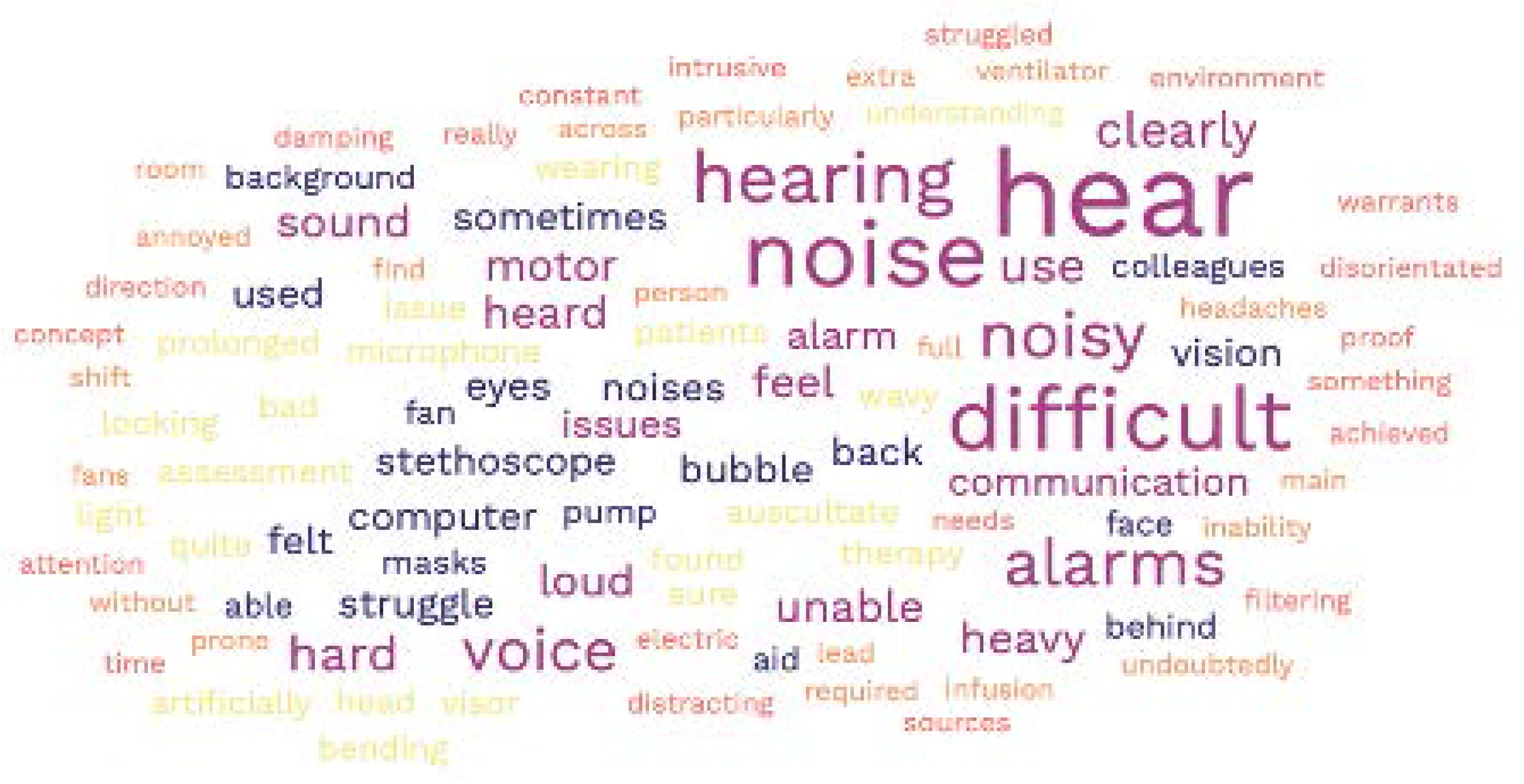
**(Supplemental)**. Word clouds from the free text feedback. Negative comments (all categories).

**Figure S6.**
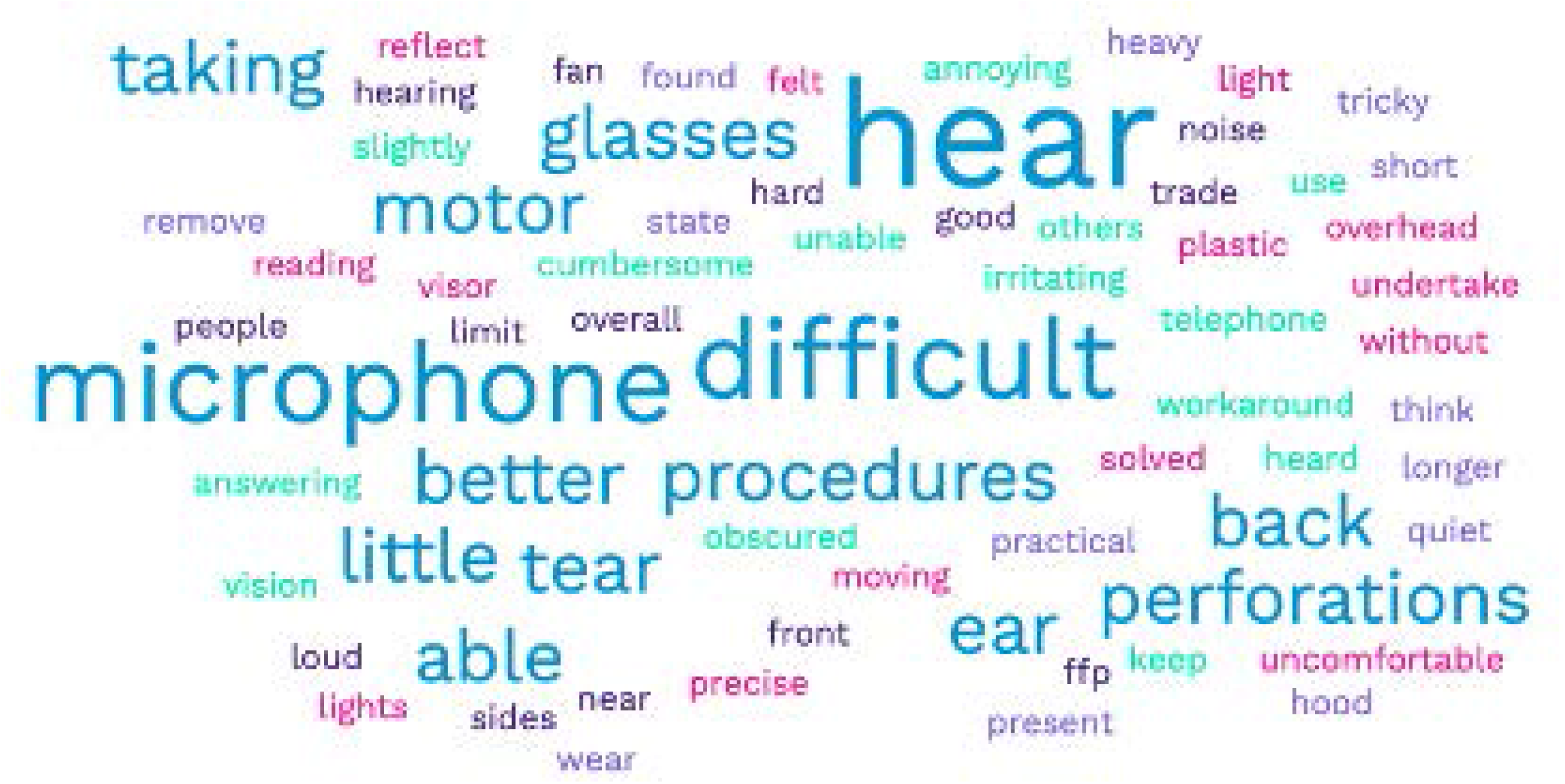
**(Supplemental)**. Word clouds from the free text feedback. Neutral comments (all categories).

**Figure S7.**
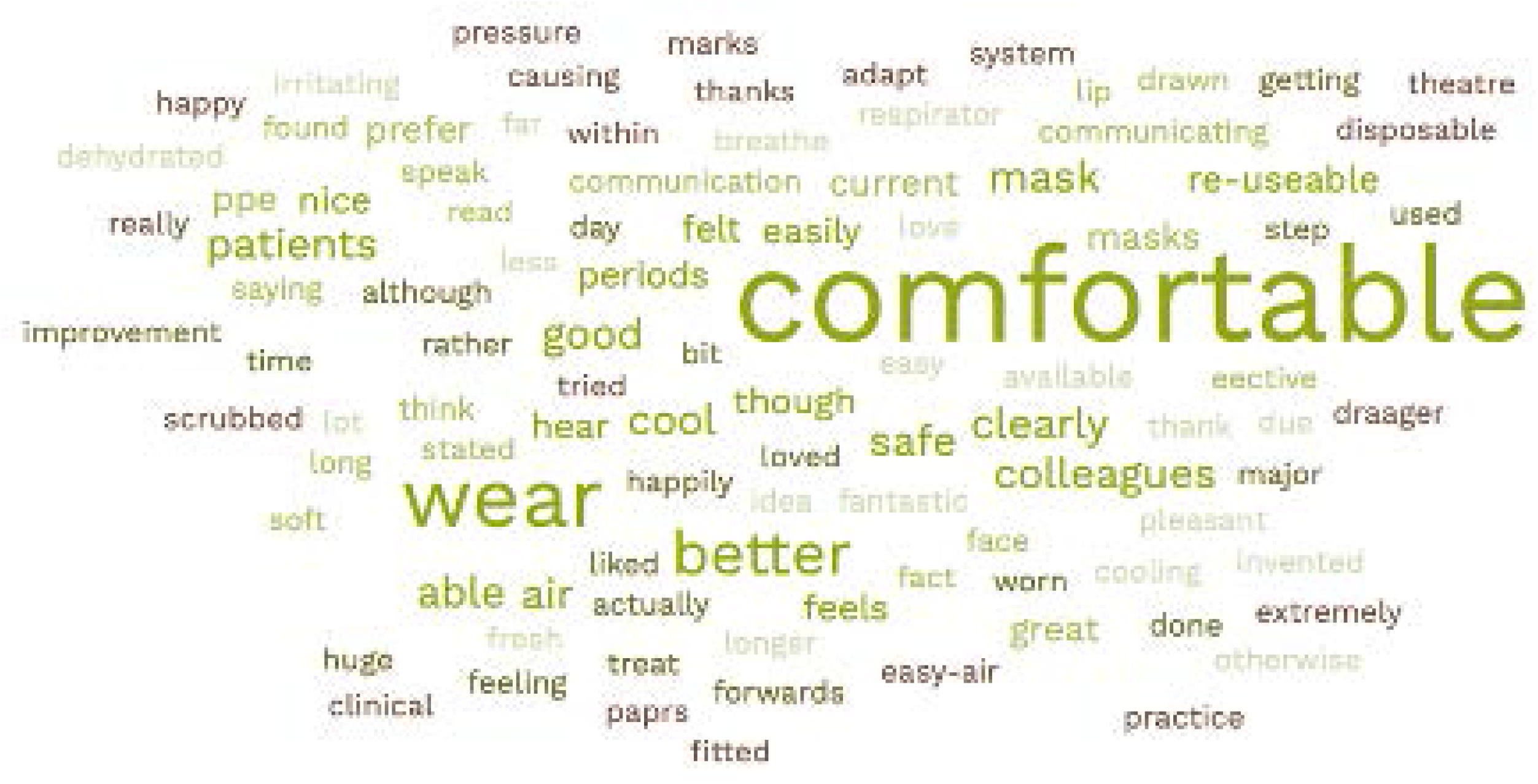
**(Supplemental)**. Word clouds from the free text feedback. Positive comments (all categories).

